# Estimating excess visual loss in people with neovascular age-related macular degeneration during the COVID-19 pandemic

**DOI:** 10.1101/2020.06.02.20120642

**Authors:** Darren S. Thomas, Alasdair N Warwick, Abraham Olvera-Barrios, Catherine Egan, Roy Schwartz, Sudeshna Patra, Haralabos Eleftheriadis, Anthony P. Khawaja, Andrew Lotery, Philipp L. Müeller, Robin Hamilton, Ella Preston, Paul Taylor, Adnan Tufail, UK EMR Users Group

## Abstract

**Objectives:** To report the reduction in new neovascular age-related macular degeneration (nAMD) referrals during the COVID-19 pandemic and estimate the impact of delayed treatment on visual outcomes at one year.

**Design:** Retrospective clinical audit and simulation model.

**Setting:** Multiple UK NHS ophthalmology centres.

**Participants:** Data on the reduction in new nAMD referrals was obtained from four NHS Trusts in England comparing April 2020 to April 2019. To estimate the potential impact on one-year visual outcomes, a stratified bootstrap simulation model was developed drawing on an electronic medical records dataset of 20,825 nAMD eyes from 27 NHS Trusts.

**Main outcome measures:** Simulated mean visual acuity and proportions of eyes with vision ≤6/60, ≤6/24 and ≥6/12 at one year under four hypothetical scenarios: no treatment delay, 3, 6 and 9-month treatment delays. Estimated additional number of eyes with vision ≤6/60 at one year nationally.

**Results:** The number of nAMD referrals at four major eye treatment hospital groups based in England dropped on average by 72% (range 65 to 87%) in April 2020 compared to April 2019. Simulated one-year visual outcomes for 1000 nAMD eyes with a 3-month treatment delay suggested an increase in the proportion of eyes with vision ≤6/60 from 15.5% (13.2 to 17.9) to 23.3% (20.7 to25.9), and a decrease in the proportion of eyes with vision ≥6/12 (driving vision) from 35.1% (32.1 to 38.1) to 26.4% (23.8 to29.2). Outcomes worsened incrementally with longer modelled delays. Assuming nAMD referrals are reduced to this level at the national level for only one month, these simulated results suggest an additional 186-365 eyes with vision ≤6/60 at one-year with even a short treatment delay.

**Conclusions:** We report a large decrease in nAMD referrals during the first month of COVID-19 lockdown and provide an important public health message regarding the risk of delayed treatment. As a conservative estimate, a treatment delay of 3 months could lead to a >50% relative increase in the number of eyes with vision ≤6/60 and 25% relative decrease in the number of eyes with driving vision at one year.

## INTRODUCTION

Healthcare services responding to the burden of the COVID-19 pandemic have had to institute policies to limit the number of patients attending hospital for other conditions. In addition, members of the public have altered their own health behaviour and reports have appeared of greatly reduced attendance at A&E and significantly lowered referral rates for some conditions.^1,2^ As the primary burden of treating COVID-19 patients has begun to ease in many countries, the implication of the epidemic for patients indirectly affected has become a focus of concern.

Ophthalmic conditions are the major source of outpatient appointments in the UK ^3^ and many, including neovascular age-related macular degeneration (nAMD) are diseases of older adults, a population at risk of developing severe COVID-19 complications.^4,5^ In England this led to a recommendation that individuals over 70 years old, the population most at risk of nAMD, should self-isolate.^6^ The Royal College of Ophthalmologists (RCOphth) produced guidance, updated on 30th March 2020, on how to triage and care for patients during the pandemic, reflecting the requirement to minimise patient contact and the associated risk for patients and staff.^7^ They advised that patients with nAMD already under review continue with all pre-planned visits and that new patients be investigated and treatment started as required. Nevertheless, as we report here, once the epidemic started, ophthalmology clinics began to experience a drop in referrals and high rates of missed appointments.

As ophthalmology clinics are now planning for the resumption of services, assessing the impact of delays to different categories of patients becomes increasingly urgent.^8^ In this paper we focus on nAMD, the leading cause of blindness in the developed world. Since patients who initiate treatment for nAMD with a better baseline visual acuity (VA), are more likely to maintain a high level of vision and achieve good long term outcomes ^9,10^ if COVID-19 delays the initiation of treatment, the outcome may have a long term impact on the burden of sight impairment and associated social independence and health consequences.

In this paper we report a change in the number of patients presenting for treatment with nAMD compared to last year and model the impact of a delay to the initiation of treatment for nAMD using a previously existing collection of data from 20,825 eyes treated in 27 different healthcare groups for nAMD.

## METHODS

### Audit of nAMD referrals

The lead clinicians for ophthalmology at multiple NHS Trusts across England were contacted to provide audit data on the number of nAMD referrals during April 2020 compared with April 2019. At the time of writing, four Trusts had responded: Moorfields Eye Hospital, King’s College London Hospital, University Hospital Southampton and Whipps Cross Hospital.

### Simulation model

#### Overview

To estimate the potential impact of delayed treatment on one-year visual outcomes, a simulation model based on a large EMR dataset of treated nAMD eyes was designed (summarised in figure 1). The key visual outcomes of interest included VA at one year and the proportions of eyes with VA ≤35 letters (6/60 Snellen), ≤55 letters (6/24+ Snellen) and ≥70 letters (6/12+ Snellen) that map approximately to the UK criteria for severe sight impairment and sight impairment (group 2), and the UK criteria for driving vision, respectively.^11,12^ These outcomes comply with recommendations from the International Consortium for Health Outcomes Measurement (ICHOM) AMD working group and were compared for four modelled scenarios: no treatment delay and 3, 6 and 9-month treatment delays (fig 1). ^13^ Of note the UK criteria for visual impairment are defined in Snellen, wheras in our UK EMR Group dataset VA is recorded in Early Treatment Diabetic Retinopathy Study (ETDRS) letters.

**Figure 1.**
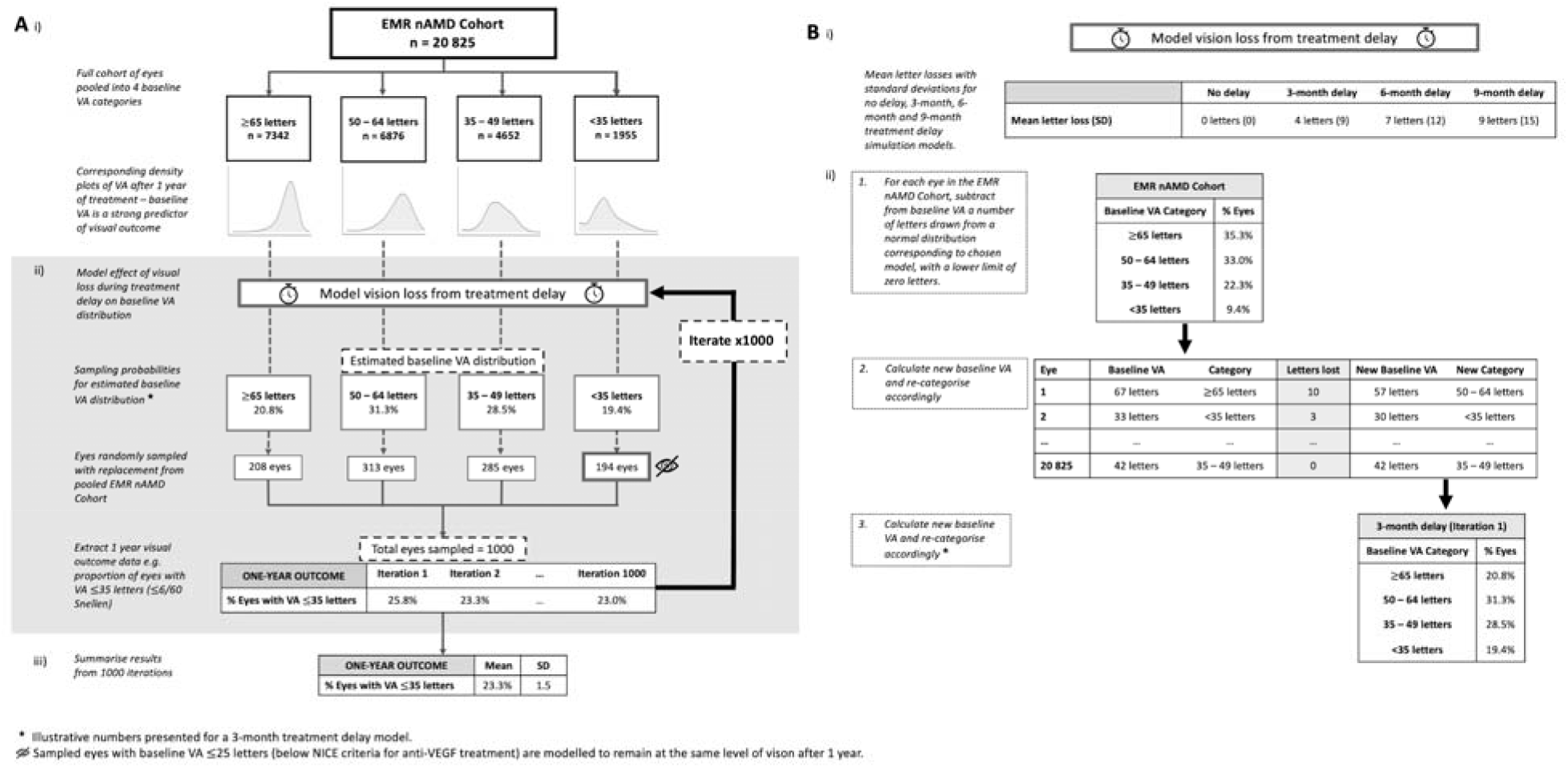
A: Summary of the simulation process. B: Modelling process for estimating the effect of vision loss during delayed treatment on baseline VA. Letter losses for the delayed treatment models are based on data from the Marina Randomised Control Trial control arm.

A recent study supported 35 letters mapping over to about 3/60 Snellen (UK criteria for severe sight impairment, group 1), and 70 letters to 6/12+ Snellen.^12^ In addition, visual field deficits may also alter visual impairment status. These factors guided our choice of ETDRS letter values for sight impairment.^11^

#### EMR data

Thirty six NHS trusts known to make comprehensive use of the Medisoft EMR system (http://www.medisoft.co.uk) to store detailed structured clinical records of patients were invited to contribute data to studies of the treatment of retinal diseases, including nAMD. Twenty-seven agreed to supply data. Patients that underwent anti-vascular endothelial growth factor (VEGF) treatment for nAMD between 2009 and 2018, and also had VA measurements recorded at first injection (baseline) and one year, were included. Patients with missing age and gender data were excluded. Patients who were not treatment naïve or were receiving one of the treatments of interest for another condition were excluded. Visual acuity measurement was performed as a part of routine clinical practice using an ETDRS chart to give an ETDRS letter score.

#### Simulation model

##### No treatment delay

The simulation process was based on a stratified bootstrapping procedure. First, all eyes in the EMR nAMD dataset were pooled into the four baseline VA bins (<35, 35-49, 50-64, ≥65 letters). Second, to simulate a range of year visual outcomes for nAMD with no treatment delay, we sampled with replacement a total of 1000 eyes from these pools with probabilities equal to the proportion of eyes in each category (table 1). One-year visual outcome measures for the sampled eyes were extracted and this process was repeated 1000 times. Finally, the range of visual outcome values generated was summarised.

**Table 1.**
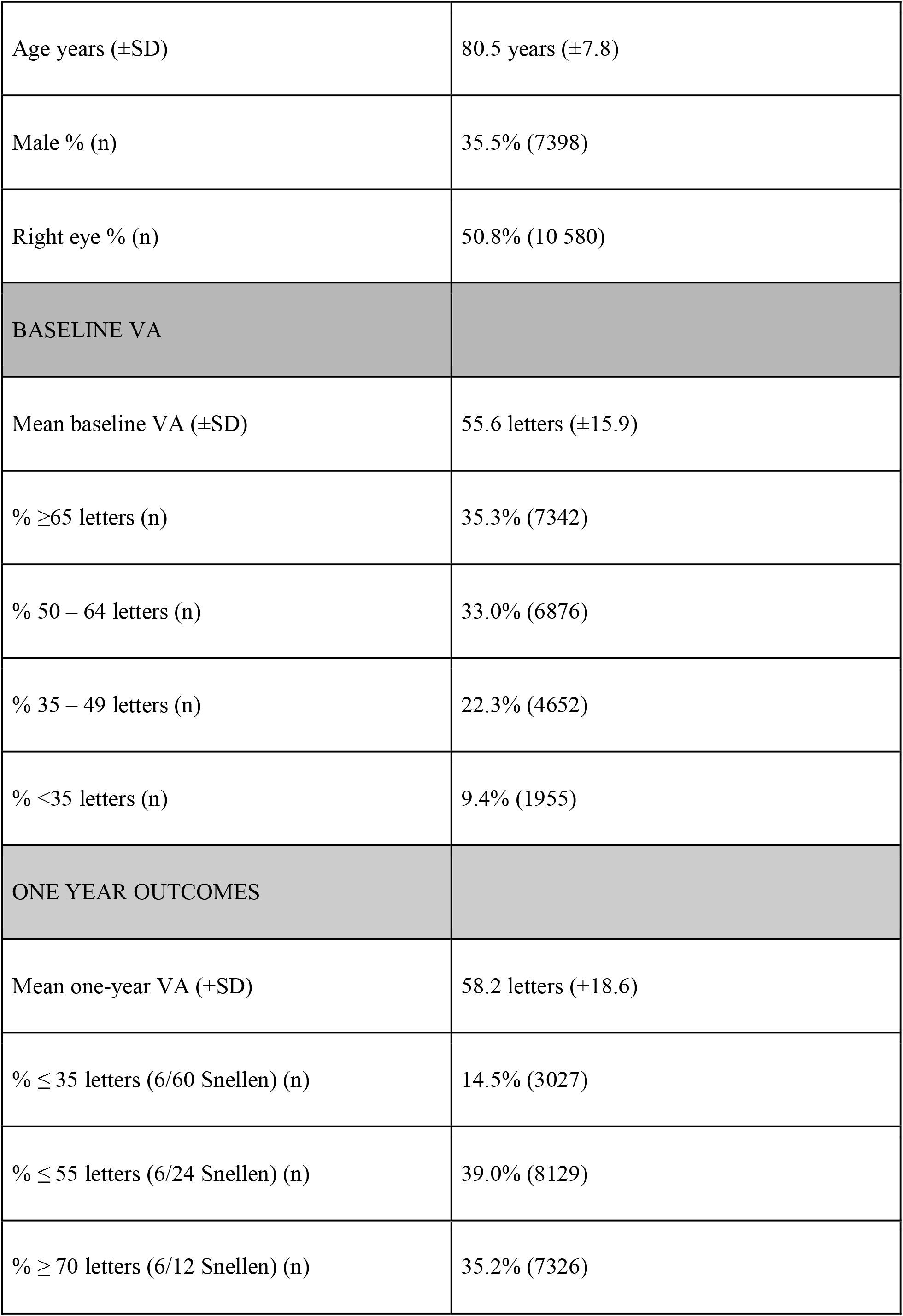
Summary of patient demographics, baseline visual acuity and one-year visual outcomes for the full EMR nAMD cohort of 20 825 eyes (18 340 patients) treated for nAMD. Abbreviations: SD - standard deviation.

|  |  |
| --- | --- |
| Age years (±SD) | 80.5 years (±7.8) |
| Male % (n) | 35.5% (7398) |
| Right eye % (n) | 50.8% (10 580) |
| **BASELINE VA** |  |
| Mean baseline VA (±SD) | 55.6 letters (±15.9) |
| % ≥65 letters (n) | 35.3% (7342) |
| % 50 – 64 letters (n) | 33.0% (6876) |
| % 35 – 49 letters (n) | 22.3% (4652) |
| % <35 letters (n) | 9.4% (1955) |
| **ONE YEAR OUTCOMES** |  |
| Mean one-year VA (±SD) | 58.2 letters (±18.6) |
| % ≤ 35 letters (6/60 Snellen) (n) | 14.5% (3027) |
| % ≤ 55 letters (6/24 Snellen) (n) | 39.0% (8129) |
| % ≥ 70 letters (6/12 Snellen) (n) | 35.2% (7326) |

##### Delayed treatment - 3, 6, and 9 months

Visual outcomes for the three delayed treatment models were estimated by varying the sampling probabilities in the simulation process described above. Baseline VA is a strong predictor of visual outcome, with good baseline VA’s tending to have better outcomes and vice versa (figure 1Ai). nAMD causes progressive visual decline and delayed treatment would therefore result in a poorer range of baseline VA’s leading to poorer visual outcomes.

To estimate the distribution of baseline VA acuities after 3, 6 and 9-month treatment delay periods, we first ascertained the approximate mean letter loss and associated standard deviations (derived from digitised standard error bars (https://automeris.io/WebPlotDigitizer)) at these time points from the MARINA Trial untreated control arm, that represents the natural history of untreated nAMD. These values are tabled in figure 1Bi. We then transformed the proportions of eyes in each baseline VA category for the no delay model in the following way. For each eye in the full cohort (n = 20 825), a randomly generated letter loss was applied to the baseline VA. The number of letters subtracted was sampled from a normal distribution based on the mean and standard deviation values obtained from the MARINA trial. We set a lower limit of zero letters loss (i.e. if a negative number of letters was sampled, then this reset to zero) as it is untreated nAMD eyes rarely improve vision.^14^ The new proportions of eyes fitting each baseline VA category were then calculated and these were applied as sampling probabilities in the simulation. This method is illustrated in figure 1Bii for a 3-month delay model. A new set of sampling probabilities was generated in this way for all 1000 iterations in each of the delayed treatment models.

Modelling by this method implicitly assumes that eyes with a given baseline VA would respond to treatment in the same way as if there had been no treatment delay. This important assumption is revisited in the discussion section.

##### Eyes with vision below NICE treatment criteria

A period of treatment delay would result in some eyes falling below standard NICE treatment criteria ≤6/96 Snellen). To reflect this in our simulation, we identified eyes with a baseline VA of ≤25 letters and modelled them to remain at the same level of VA at one year, as if they had not received treatment. This rule was applied to all models, including the no treatment delay model, to allow a fair comparison of results. The simulation was also performed without this additional rule as a sensitivity analysis (supplementary materials).

### National estimates of delayed nAMD referrals

The reduction in new nAMD referrals audited at the four aforementioned NHS Trusts was projected to the whole of the UK and results from the simulation model were then applied to estimate the potential national impact of delayed nAMD treatment.

### Statistical analysis

All analyses were implemented in R version 3.6.2.^15^ Graphs were generated using the ggplot2 package. All analysis code is available at https://github.com/rmgpanw/nAMD_tx_delay_simulation. Visual acuities are presented in ETDRS letter format unless stated otherwise.

### Patient and public involvement statement

Given the urgency of the COVID-19 situation, there were no funds or time allocated for PPI and we were unable to involve patients. We have invited the patient support group the Macular Society, and patients associated with this society to help us develop our dissemination strategy.

## RESULTS

### New nAMD referrals during the COVID-19 pandemic

Figures from the first four hospitals that responded to our request for data showed a mean drop of 72% (range 65 to 87%) in the number of referrals for nAMD in April 2020 (40 per month) compared to April 2019 142 per month).

### Simulation model

#### EMR nAMD cohort

From the EMR dataset we identified 20 825 eyes (18 340 patients) that underwent anti-VEGF treatment for nAMD between 2009 and 2018 and had VA measurements recorded at first injection (baseline) and one year. The demographics, baseline VA distribution and one-year visual outcomes for this cohort are described in table 1.

#### Simulated visual outcomes at 1 year

The average simulated mean baseline VA’s for the no delay, 3, 6 and 9-month delay models were 55.6 letters (±0.6 SD), 49.9 letters (±0.6 SD), 47.2 letters (±0.6 SD) and 45.5 letters (±0.6 SD) respectively. Figure 2 summarises the average baseline and one-year VA distributions generated for each model, as well as those for the full EMR nAMD cohort. Results for the no delay model resemble those of the whole cohort, whereas both the baseline and one-year VA distributions worsen incrementally with increasing treatment delay.

**Figure 2.**
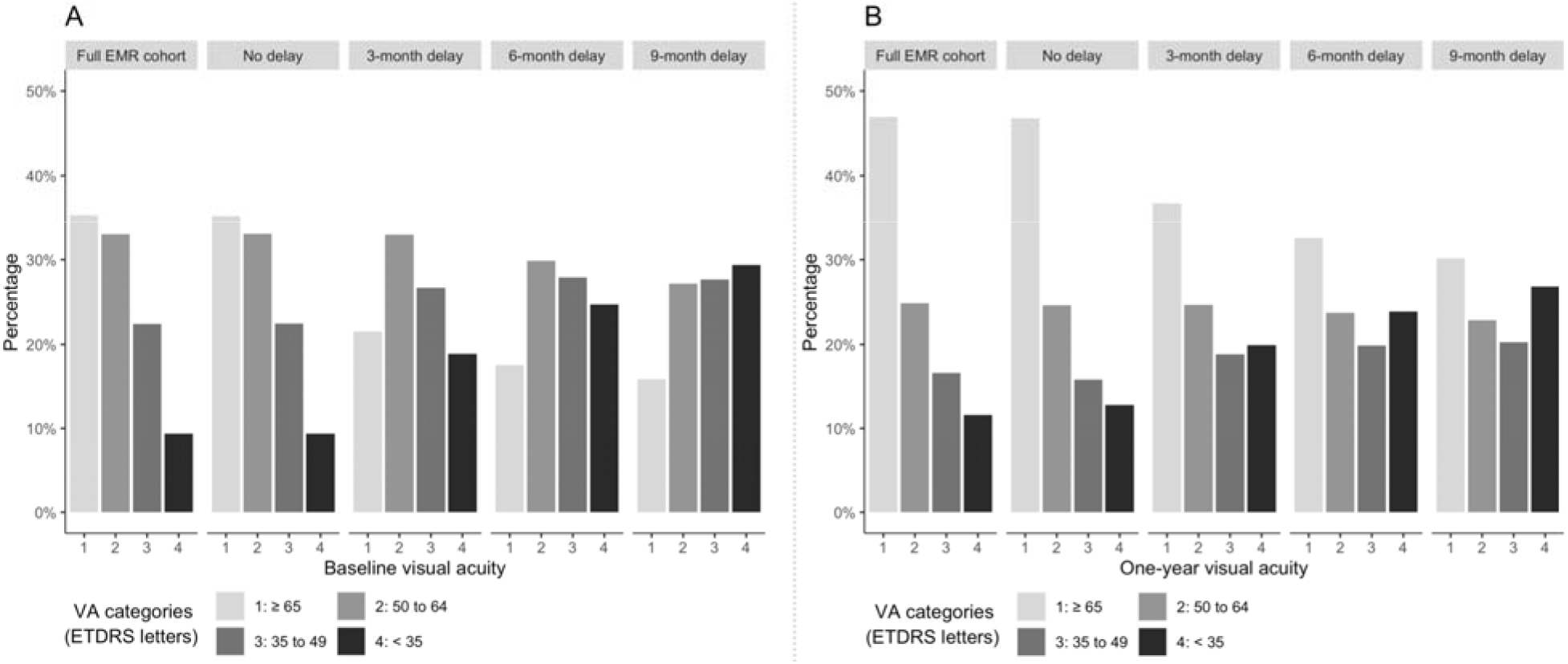
The average distribution of visual acuities across all iterations in the simulation process at baseline (A) and at one year (B) for the full EMR nAMD cohort and under four modelled conditions: no, 3, 6 and 9-months treatment delay.

Average simulated one-year visual outcomes for the four models are presented in table 2. These results are complemented by figure 3 which graphically depicts the range of estimates generated across 1000 iterations for each model, showing minimal overlap between the no delay model with any of the treatment delay models.

**Table 2.** Average (95% bootstrap confidence interval) simulated one-year visual outcomes under each of the four modelled conditions

|  | No treatment delay | 3-month delay | 6-month delay | 9-month delay |
| --- | --- | --- | --- | --- |
| **Mean one-year VA** | 57.8 letters<br>(56.6 - 59.1) | 52.7 letters<br>(51.4 - 54.1) | 50.3 letters<br>(48.8 - 51.6) | 48.6 letters<br>(47.2 - 49.9) |
| **% $\leq 35$ letters** | 15.5%<br>(13.2 - 17.9) | 23.3%<br>(20.7 - 25.9) | 27.6%<br>(24.7 - 30.6) | 30.8%<br>(28.0 - 33.7) |
| **% $\leq 55$ letters** | 39.4%<br>(36.1 - 42.5) | 50.0%<br>(46.9 - 53.1) | 54.8%<br>(51.5 - 58.2) | 57.9%<br>(54.7 - 61.1) |
| **% $\geq 70$** | 35.1% | 26.4% | 23.1% | 21.3% |
| **letters** | (32.1 - 38.1) | (23.8 - 29.2) | (20.6 - 25.7) | (18.7 - 23.9) |

**Figure 3.**
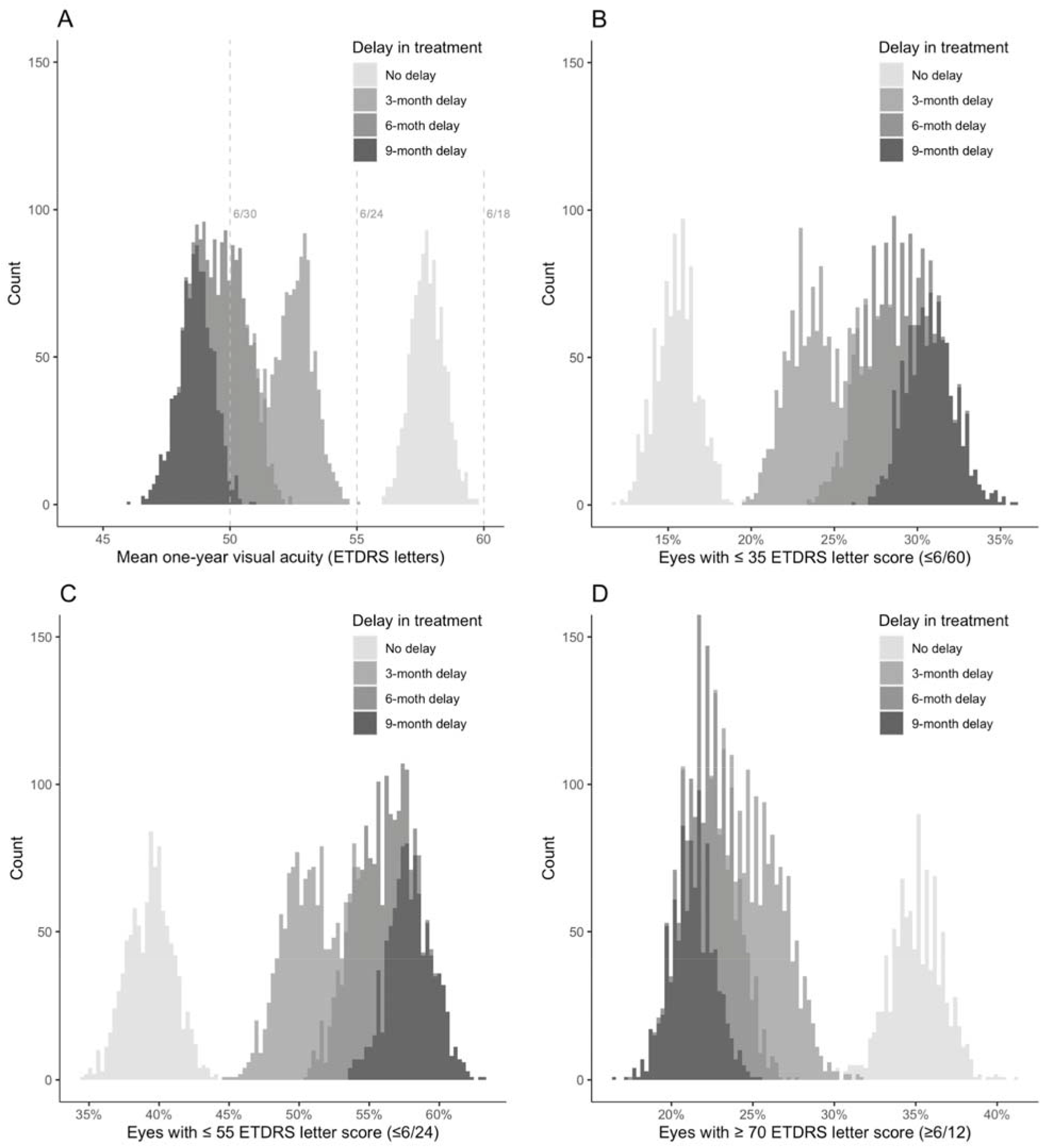
Overlaid histograms showing the range of simulated one-year visual acuity (VA) outcomes across 1000 iterations for the no delay, 3, 6 and 9-month treatment delay models. ‘Count’ on the y-axis refers to the number of iterations that returned a visual outcome estimate at the values on the x-axis. A: mean one-year VA (dashed grey vertical lines illustrate Snellen equivalents). B: percentage of eyes with ≤35 letters (≤6/60 Snellen). C: percentage of eyes with ≤55 letters (≤6/24+ Snellen) D: percentage of eyes with ≥70 letters (≥6/12+ Snellen).

### Estimated national impact of delayed nAMD treatment

Owen, Jarrar, Wormold et al. estimate the annual incidence of nAMD to be 39,700.^16^ If the COVID19 pandemic affects patient referrals for 3 months, and over that period they are reduced by 72%, 7146 patients will delay initiating treatment. We estimate that if these patients did not delay treatment then 1108 eyes would have a VA ≤35 letters (severe sight impairment or legal blindness) after 12 months. This rises to between 1887 and 2201 if treatment initiation is delayed by 3 to 9 months. The number of people who would meet the legal driving limit would be 2508 (27.8% of the total) with no delay but fall to between 1887 and 1522 under the different modelled delays. If referrals are reduced only for one month, the figures will be one third of those quoted, but this will still estimate between 186 and 365 excess eyes becoming legally blind at one year.

## DISCUSSION

During the COVID-19 lockdown people over 70 years old have been told to self-isolate. We report a 72% reduction in the expected number of referrals for nAMD at four UK nAMD treatment centres, suggesting there is a substantial number of patients with new nAMD who will suffer from delayed treatment. We also estimate the potential impact of this by simulating one-year visual outcomes following various periods of treatment delay and projecting these results to a national level. It has been well documented that AMD negatively affects daily living tasks including mobility, face recognition, computer use, meal preparation, cleaning, watching TV, reading, driving and, in some cases, self-care.^17^ The consequences of untreated nAMD are likely to exacerbate the problems of COVID-19 and non-COVID-19 induced social isolation on the mental and physical health of the elderly.^18^

The aim of our simulation model was not to provide an absolute estimate of the additional burden of VA loss from delayed nAMD treatment during the COVID-19 pandemic, but rather to provide conservative estimates with some indication of variability due to chance. The simulation is based on a large dataset of 20 825 eyes treated at sites across the UK over the past 10 years. It is reasonable therefore to assume that the range of presentations and visual outcomes is representative of the UK population as a whole. In reality, the presenting characteristics and visual outcomes of nAMD cases will fluctuate randomly between shorter time periods. We mirror this by adopting a stratified bootstrapping approach with 1000 iterations for each simulation model, predicting the degree of possible variation around average visual outcome estimates.

The method used to estimate vision loss during a period of delayed treatment was based on high quality data from the pivotal MARINA trial control arm.^14^ This represents the best available natural history data for nAMD. A lower limit of zero letters lost was applied during this process since it is improbable for untreated nAMD to result in improved VA. The average differences in mean baseline VA between the no delay and 3, 6 and 9-month delay models (5.7, 8.4 and 10.1 letters respectively) were therefore inflated slightly above the corresponding mean letter losses reported in MARINA (approximately 4, 7 and 9 letters respectively). However, the mean baseline VA in the MARINA trial (53.6 letters) was lower than that of the EMR cohort (55.6 letters) - while a better baseline VA is predictive of better visual outcomes, it also carries an increased chance of higher degrees of letter loss. This suggests that the hypothetical mean letter losses for this EMR cohort at 3, 6 and 9 months if they had been left untreated are indeed likely to have been higher than those reported in MARINA.

The EMR dataset includes 725 treated eyes (3.5% of the full EMR nAMD cohort) with a baseline VA of ≤25 letters (≤6/96 Snellen), which falls outside standard NICE treatment criteria. The decision to treat in these cases may have been made either on compassionate grounds or based on VA measurements that fell within NICE treatment criteria at the time of listing for treatment. These eyes were modelled to remain at the same level of vision at one year, as if they had not received treatment. We believe this more accurately represents reality, as visual loss during a period of treatment delay would result in a higher proportion of new nAMD eyes falling below NICE treatment criteria by the time they reached clinic. This rule was also applied to the no delay model to allow fair comparison with delayed treatment scenarios. Despite this, visual outcome results from the no delay scenario still closely resembled those for the whole EMR cohort. The simulation was also run without applying this rule as a sensitivity analysis (Supplementary materials - sFigure 1, sTable 1 and sFigure 2). This still returned similar, albeit slightly reduced, estimated differences in visual outcomes between the no treatment delay and treatment delay models.

A key assumption underlying the treatment delay models is that eyes receiving treatment late in the course of the disease will respond as well as if there had been no treatment delay. This underestimates the true burden of additional visual loss from treatment delay as untreated nAMD causes retinal scarring, which would lead to progressively worse treatment responses with longer delays. It follows that the degree to which our simulation process underestimates visual loss would also therefore increase with longer modelled delays. Overall, the presented estimates for visual loss incurred by a period of treatment delay are almost certainly underestimates of the true values.

Following guidelines from The Royal College of Ophthalmologists, treating centres for nAMD have endeavoured to continue treating this condition for patients already diagnosed, and delaying follow up of other less urgent conditions to allow capacity for safe spacing for this high COVID- 19 risk group.^19^ Despite these interventions, many patients (between 5 and 25%) have not attended their appointment in the four centres surveyed. What is of more concern is the large drop in patients presenting with new nAMD, which, according to data from four large treating centres, is about 72% less than expected. Projecting this nationally, using an accepted estimate of the incidence rate of nAMD then 2388 patients will have delayed treatment in the month of April alone (2123-2620 95% confidence interval). We estimate that this will lead to between 234 and 470 additional cases of legal blindness for nAMD, if the partner eye was already below the threshold. However, this is a conservative estimate, derived from a one-month reduction in presentation with a 3-month delay before treatment and assuming that treatment, once initiated on the delayed eyes, has a similar benefit as when initiated promptly. If patients with other sight-threatening diseases behave in a similar manner to those studied here, the increase in numbers of patients with preventable legal blindness might be distinctly higher.

In summary, adopting a conservative model our estimates still indicate a substantial increase in visual loss (legal blindness, loss of driving vision) from delayed nAMD treatment, lending strong support to an important public health message. Isolating the elderly might reduce transmission, however this will lead to visual loss in those at risk of nAMD, predominantly elderly women. From a purely ocular standpoint, It is therefore imperative that those at risk are advised to seek care in a timely fashion, before irreversible vision loss occurs. Equally and more holistically, safe transport and socially-distanced diagnostic and treatment environments must be provided and instructions on how to access these resources communicated effectively to patients, GPs, optometrists and patient support groups. A continuing reduction in patients presenting for treatment will add to the cumulative indirect healthcare burden that COVID-19 is already having on health, well-being and social care costs. Public Health messaging via national and local agencies and patient support groups may be warranted to allow for timely treatment to prevent visual loss.

## Data Availability

Data availability statement. All analysis code is available

